# Oral Gene III^®^ L-Ergothioneine Capsules in Women with Suboptimal Ovarian Reserve: A Single-Center, Open-Label, Self-Controlled Pilot Study of Ovarian Reserve Markers, Endocrine Profiles, and Reproductive Aging-Related Symptoms

**DOI:** 10.64898/2026.04.02.26350093

**Authors:** Wei Liu, Cong Guo, Wei Ding, Juan Cao, Hongying Ju, Fengjuan Liu, Guohua Xiao

## Abstract

**Purpose:** To evaluate the efficacy and safety of oral L-ergothioneine (EGT) in improving ovarian reserve and clinical symptoms in women with suboptimal ovarian function. As a proof-of-concept study, we explored correlations between hormonal shifts and symptom amelioration.

**Methods:** This single-center, open-label trial enrolled 40 women (aged 35–45 years) experiencing age-related reproductive decline (baseline AMH: 1.0–3.0 ng/mL) and menstrual disorders. Participants received oral EGT (120 mg/day) for three consecutive menstrual cycles. The primary outcome was the change in serum AMH. Secondary outcomes included sex hormones (FSH, E2), antral follicle count, and validated questionnaires (KI, PSQI, SF-36) and an exploratory Menstrual Symptom Score.

**Results:** Thirty-six participants completed the intervention without product-related adverse events. EGT supplementation was associated with increases in core ovarian markers: mean AMH increased from 1.81 ± 0.72 to 2.46 ± 1.54 ng/mL (mean change +0.65 ng/mL, 95% CI [0.14, 1.17], p = 0.018). Concurrently, basal FSH decreased (8.38 ± 2.83 to 7.05 ± 2.51 mIU/mL, mean change −1.33, 95% CI [−2.50, −0.17]; p = 0.032, FDR-adjusted p = 0.048) and E2 increased (43.78 ± 18.87 to 63.46 ± 50.81 pg/mL; mean change +19.69, 95% CI [3.99, 35.38]; p = 0.019, FDR-adjusted p = 0.048). Clinical assessments showed progressive reductions in KI (5.42 ± 3.66 to 1.90 ± 2.16, p < 0.0001) and PSQI scores (6.89 ± 1.82 to 5.50 ± 1.40, p < 0.0001), alongside improved menstrual and SF-36 scores (p < 0.001). Subgroup analysis stratified by baseline ovarian reserve showed a significant AMH increase in the low-reserve subgroup (p = 0.017) but not the high-reserve subgroup. Exploratory correlation analysis showed that ΔFSH was associated with improvements in sleep quality (ΔPSQI, r = 0.43, p < 0.05) and E2 increases (r = −0.46, p < 0.05), linking hormonal stabilization directly to systemic relief.

**Conclusion:** In this open-label, single-arm pilot study, oral EGT supplementation was associated with increases in serum AMH and favorable shifts in the basal FSH/E2 profile, alongside improvements in reproductive aging-related and sleep symptoms. Because the design lacks a control group, these changes cannot be attributed to EGT alone and may partly reflect natural variation, regression to the mean, or placebo effects. These hypothesis-generating findings warrant confirmation in adequately powered, placebo-controlled trials.

**Trial Registration:** ChiCTR2500104484; Prospectively registered on 2025-06-18.

## 1. Introduction

Ovarian aging is the primary hallmark of female reproductive senescence, clinically characterized by a progressive decline in ovarian reserve, diminished oocyte quality, and dysregulation of the hypothalamic-pituitary-ovarian (HPO) axis [1]. In recent decades, environmental stressors and lifestyle shifts have contributed to an increased incidence of suboptimal ovarian reserve among younger populations. Beyond compromised fertility, suboptimal ovarian reserve precipitates early estrogen withdrawal, triggering menstrual irregularities and peri-menopausal vasomotor symptoms (e.g., hot flashes, night sweats), which severely impair physical and psychological well-being.

It is important to distinguish among related but non-equivalent concepts. Diminished or suboptimal ovarian reserve is a quantitative, biochemical concept reflecting a reduced remaining follicle pool (indexed by AMH, FSH, and antral follicle count). In contrast, the menopausal transition and perimenopause are clinically defined reproductive stages characterized by changes in menstrual cyclicity and endocrine milieu, as formalized by the STRAW+10 staging system [2]. A serum AMH of 1.0-3.0 ng/mL indicates reduced ovarian reserve but does not by itself define a perimenopausal stage. Accordingly, the present study targets women with suboptimal ovarian reserve, and the associated symptoms are referred to as reproductive aging-related symptoms rather than menopausal symptoms.

Accumulating molecular evidence indicates that oxidative stress (OS) and mitochondrial dysfunction act as primary pathological drivers of accelerated ovarian aging [3]. The physiological processes of ovulation and luteinization intrinsically generate reactive oxygen species (ROS). As the endogenous antioxidant defense systems within granulosa cells and oocytes deteriorate with age, ROS overaccumulation leads to lipid peroxidation, mitochondrial DNA damage, and accelerated apoptosis of granulosa cells [4, 5]. This oxidative microenvironment hastens the depletion of the primordial follicle pool, clinically reflecting as an irreversible drop in anti-Müllerian hormone (AMH) levels [6]. Consequently, targeting follicular OS with targeted, high-efficiency antioxidants represents a promising strategy for preserving ovarian function.

L-ergothioneine (EGT) is a naturally occurring, highly stable sulfur-containing amino acid derivative synthesized predominantly by fungi and mycobacteria [7]. Distinct from classical antioxidants, EGT utilizes a highly specific organic cation transporter, OCTN1 (encoded by the *SLC22A4* gene), to cross cell membranes [8]. Because OCTN1 is highly expressed in tissues subjected to intense oxidative stress, EGT actively accumulates within mitochondria, where it robustly scavenges free radicals, protects DNA from oxidative damage, and mitigates cellular senescence [9, 10]. EGT’s excellent safety profile has been extensively validated in toxicological studies. Notably, the high-purity EGT has been granted Generally Recognized as Safe (GRAS) status by the U.S. Food and Drug Administration (FDA, GRN No. 1270), alongside safety validations by the European Food Safety Authority (EFSA) [11–13].

Despite its established safety profile and cytoprotective properties [14], clinical data regarding the therapeutic application of EGT for female reproductive health and ovarian reserve modulation remain highly limited. Therefore, this clinical trial was designed to evaluate the efficacy and safety of oral EGT capsules in modulating serum AMH, optimizing endocrine profiles, and alleviating associated systemic symptoms in women experiencing age-related or stress-induced sub-optimal ovarian function.

## 2. Methods

### 2.1 Study Design

This prospective, single-center, open-label, self-controlled clinical trial was conducted at the Qingdao Central Hospital of Rehabilitation University between September 2025 and February 2026. The study protocol complied with the Declaration of Helsinki and Good Clinical Practice (GCP) guidelines and was formally approved by the Institutional Ethics Committee of Qingdao Central Hospital (Approval No.: SY202502202). Written informed consent was obtained from all participants prior to screening.

Although the clinical trial was initially registered with The First People’s Hospital of Hefei, the clinical execution and participant enrollment were subsequently transferred to Qingdao Central Hospital due to logistical considerations. The ethics committee of the final executing site formally approved all protocols.

As this was an exploratory, proof-of-concept pilot study evaluating the novel application of EGT in ovarian aging, the sample size was not strictly based on a formal power calculation. A target enrollment of 40 participants was determined based on clinical feasibility and established norms for preliminary dietary supplement interventions, aimed at providing effect size estimates for powering future large-scale, randomized controlled trials.

### 2.2 Participants

The study enrolled 40 female participants based on stringent criteria.

Eligible participants were women aged 35-45 years with menstrual disorders (e.g., oligomenorrhea, polymenorrhea, altered cycle length, abnormal menstrual volume, or dysmenorrhea) and biochemically confirmed suboptimal ovarian reserve (serum AMH 1.0-3.0 ng/mL), who provided written informed consent. Women were excluded if they had severe cardiovascular, hepatic, or renal disease; a history of psychiatric disorders; oophorectomy or surgery affecting ovarian reserve; polycystic ovary syndrome, hyperprolactinemia, hyperandrogenism, diabetes, or thyroid dysfunction; were pregnant or lactating; had a history of substance abuse; or had used hormone preparations or other antioxidant supplements within the past month.

Participant flow and study adherence were systematically tracked and visualized (Figure 1). Baseline demographic characteristics, including age, reproductive history, occupational stress, and baseline clinical parameters, were collected and summarized to assess cohort representativeness (Table 1).

**Figure 1.**
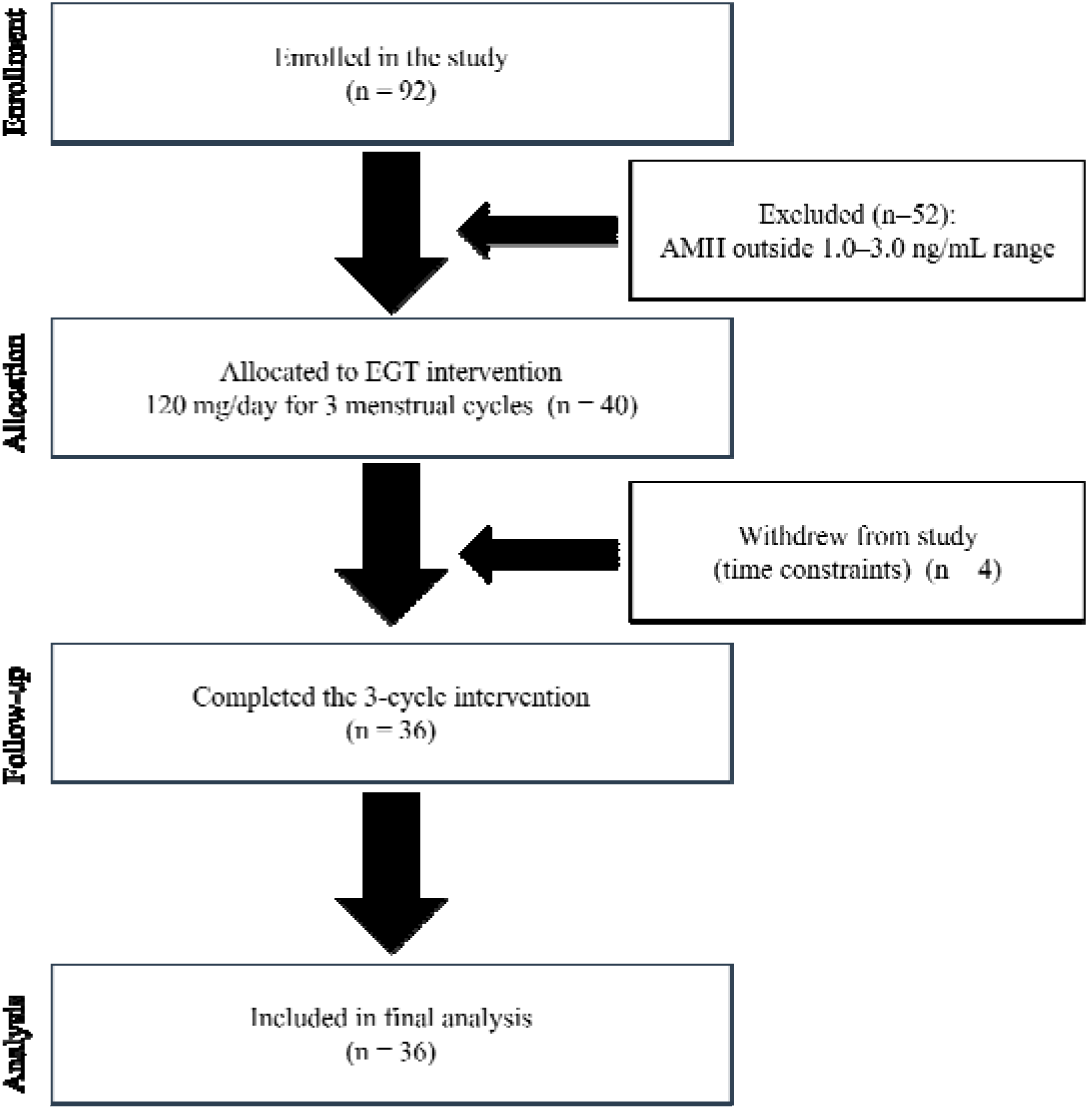
CONSORT flow diagram of the study. 92 women assessed for eligibility, 52 were excluded because their serum AMH fell outside the 1.0–3.0 ng/mL range. Forty women were enrolled and allocated to oral L-ergothioneine (120 mg/day); 4 withdrew due to time constraints, and 36 completed the 3-cycle intervention and were included in the final analysis.

**Table 1.** Baseline Demographic and Clinical Characteristics of the Analyzed Participants (N = 36)

| Characteristics | Values (Mean ± SD) |
| --- | --- |
| **Demographics** |  |
| Age (years) | 39.81 ± 2.92 |
| Body Mass Index (BMI, kg/m<sup>2</sup>) | 23.25 ± 2.53 |
| **Objective Ovarian & Endocrine Markers** |  |
| Baseline AMH (ng/mL) | 1.81 ± 0.72 |
| Baseline FSH (mIU/mL) | 8.38 ± 2.83 |
| Baseline Estradiol (E2, pg/mL) | 43.78 ± 18.87 |
| Baseline LH (mIU/mL) | 5.87 ± 2.12 |
| Baseline Antral Follicle Count (AFC) | 10.1 ± 2.6 |
| **Subjective Clinical Scores** |  |
| Baseline Modified Kupperman Index | 5.42 ± 3.66 |
| Baseline PSQI Total Score | 6.89 ± 1.82 |
| Baseline SF-36 Score | 134.39 ± 1.50 |
| Data are expressed as mean ± SD for continuous variables. | |

### 2.3 Interventions

Eligible participants received Gene III^®^ L-ergothioneine capsules (30 mg EGT per capsule; Batch No. P776678), administered as a dietary antioxidant supplement, supplied by Jiangsu Gene III Biotechnology Co., Ltd. The prescribed dosage was 120 mg/day, administered as two capsules in the morning and two capsules in the evening. The intervention spanned three consecutive menstrual cycles. Concomitant use of any other medications or nutritional supplements capable of altering ovarian function was strictly prohibited throughout the study period.

Crucially, the proprietary EGT ingredient utilized in this trial has received a “No Questions” GRAS designation from the U.S. FDA (GRN No. 1270). The 120 mg/day intervention dosage was established well within the broad safety margins derived from the comprehensive toxicological evaluations supporting this GRAS status, ensuring high clinical tolerability.

### 2.4 Outcome Measures

Outcomes were pre-specified in a four-tier hierarchy:

Primary endpoint: change in serum AMH from baseline to V3.

Secondary endpoints: changes in basal FSH, E2, and antral follicle count (AFC).

Exploratory endpoints: other hormones (LH, PRL, P, T), ovarian volume, modified Kupperman Index (KI) [15], PSQI [16], SF-36 [17], and the Menstrual Status score.

Safety endpoints: adverse events, vital signs, complete blood count, urinalysis, and liver/kidney function.

Menstrual Status Questionnaire: This questionnaire was developed for this study and has not been formally validated; therefore, menstrual score findings should be interpreted as exploratory. It comprised five domains—cycle regularity, menstrual duration, menstrual volume, dysmenorrhea, and cycle predictability—each scored on an ordinal scale (0, 3, or 5 points depending on the domain). The five domain scores were summed to yield a total score ranging from 0 to 25, with higher scores indicating better menstrual health. Amenorrhea or markedly irregular cycles (variation >7 days) were scored 0 within the cycle-regularity domain. The English version of the questionnaire is provided in Fig. S2.

### 2.5 Statistical Analysis

Statistical analyses were performed using SAS software, version 9.4. Continuous variables were presented as mean ± standard deviation (SD). Intra-group comparisons between baseline and post-treatment time points were evaluated using paired t-tests for normally distributed data, or the Wilcoxon signed-rank test for non-parametric data. A two-sided P < 0.05 was considered statistically significant. The primary endpoint (AMH) was tested without multiplicity adjustment. For the three secondary endpoints (FSH, E2, AFC), the Benjamini-Hochberg false discovery rate (FDR) procedure was applied, and adjusted p-values are reported. All other outcomes were exploratory and interpreted as hypothesis-generating, without formal correction. Mean changes from baseline are reported with 95% confidence intervals. Analyses used the per-protocol population (N = 36).

In addition to pre-post comparisons, exploratory subgroup analyses were conducted by stratifying participants according to baseline ovarian reserve (median split of baseline AMH), a biologically grounded stratification more directly reflecting follicular status than chronological age. To evaluate the relationship between endocrine markers (AMH, FSH, E2) and clinical symptoms (KI, PSQI, SF-36), Pearson or Spearman correlation coefficients were calculated based on the change from baseline (Δvalues) to V3. Responders were defined a priori as participants exhibiting a ≥10% increase in baseline AMH, and baseline characteristics were compared between responders and non-responders.

## 3. Results

### 3.1 Study Adherence and Participant Demographics

A total of 92 female candidates were initially screened for eligibility. Among them, 56 were excluded based on the inclusion/exclusion criteria, resulting in an enrolled cohort of 40 participants with diminished ovarian reserve (DOR) who were allocated to the single-arm EGT intervention. Following allocation, four participants voluntarily withdrew prior to completion due to personal time constraints. Ultimately, 36 participants (90% adherence rate) successfully completed the continuous 3-cycle intervention and were included in the final statistical analysis (Figure 1). The baseline demographic and clinical characteristics of the analyzed cohort are summarized in Table 1. The consistency across subjective scores (KI, PSQI) and objective hormones (AMH, FSH) at baseline indicated a relatively homogeneous state of reproductive decline prior to intervention.

### 3.2 Changes in Objective Ovarian Reserve Markers

The primary clinical endpoint, serum AMH levels, demonstrated a statistically significant recovery following the 3-month EGT intervention. Mean serum AMH increased from 1.81± 0.72 ng/mL at V0 to 2.46 ± 1.54 ng/mL (mean change +0.65 ng/mL, 95% CI [0.14, 1.17]) at V3 (p = 0.018). The individual trajectories overlaying the violin distribution reveal a clear upward trend for a substantial majority of participants (Figure 2A). Concurrently, basal FSH levels decreased from 8.38 ± 2.83 mIU/mL to 7.05 ± 2.51 mIU/mL (mean change −1.33, 95% CI [−2.50, −0.17]; FDR-adjusted p = 0.048) (Figure 2B). Crucially, this was accompanied by a supportive increase in estradiol (E2) levels, rising from 43.78 ± 18.87 pg/mL to 63.46 ± 50.81 pg/mL (mean change+19.69, 95% CI [3.99, 35.38]; FDR-adjusted p = 0.048) (Figure 2C). Exploratory secondary markers, including other basal sex hormones (LH, PRL, P, T) and ovarian volume, remained stable throughout the observation period (Supplementary Table S1). Antral follicle count (AFC) showed a non-significant numerical increase from 10.1 ± 2.6 at V0 to 10.6 ± 3.2 at V3 (FDR-adjusted p = 0.419), and no conclusion regarding EGT’s effect on follicle count can be drawn within this timeframe. Individual trajectories are shown in Supplementary Figure S1.

**Figure 2.**
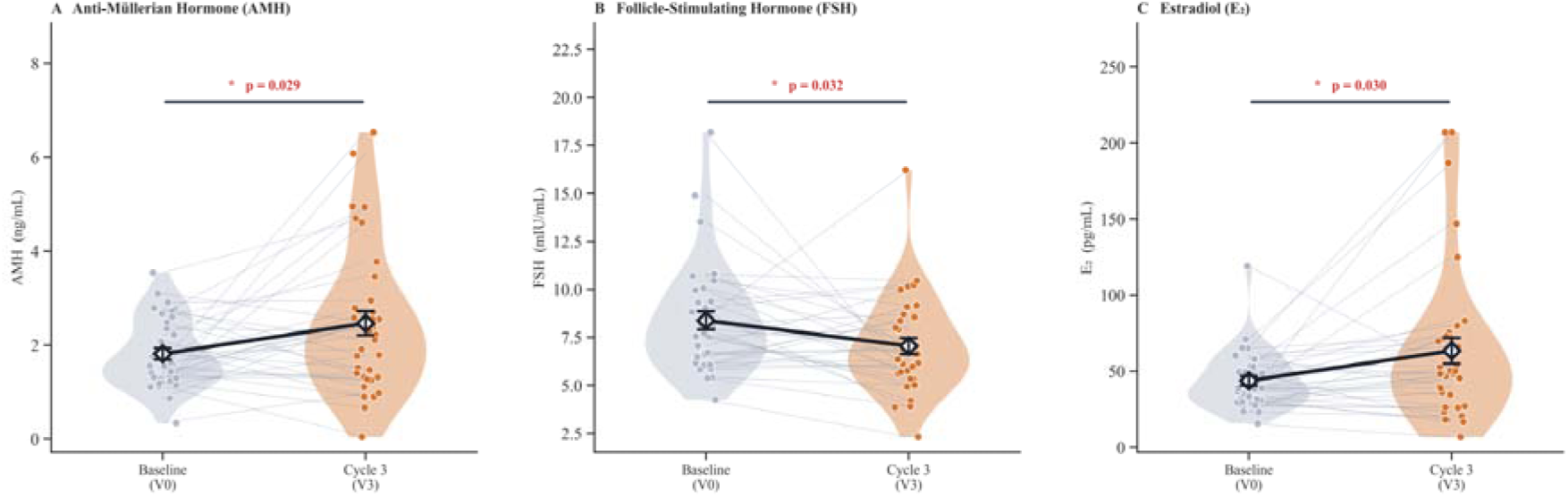
Individual trajectories and distribution of ovarian reserve and endocrine markers. Violin plots show serum levels at baseline (V0) and after three menstrual cycles (V3) for (A) AMH, (B) FSH, and (C) E2. Overlaid spaghetti lines represent paired individual trajectories (N =36); solid black lines indicate group means. *P < 0.05 vs. baseline.

### 3.3 Depth of Analysis: Subgroup Heterogeneity and Mechanisms of Symptom Relief

We conducted two additional exploratory analyses to characterize response heterogeneity and hormonal–symptom relationships:

Subgroup Analysis by Baseline Ovarian Reserve: To explore whether the magnitude of AMH response varied by pre-treatment follicular status, participants were stratified by a median split of baseline AMH (1.60 ng/mL; n=18 per subgroup). In the low-reserve subgroup (AMH ≤1.60 ng/mL), AMH increased from 1.25 ± 0.30 to 2.24 ± 1.58 ng/mL (mean change +0.99 ng/mL, 95% CI [0.26, 1.72]; p = 0.017), representing a clinically meaningful response. In contrast, the high-reserve subgroup (AMH >1.60 ng/mL) showed a smaller, non-significant increase (+0.31 ng/mL, 95% CI [−0.40, 1.02]; p = 0.402) (Figure 3A). This differential response suggests that women with poorer baseline ovarian reserve may derive greater benefit from EGT supplementation, consistent with the greater scope for improvement in a more compromised follicular microenvironment.

**Figure 3.**
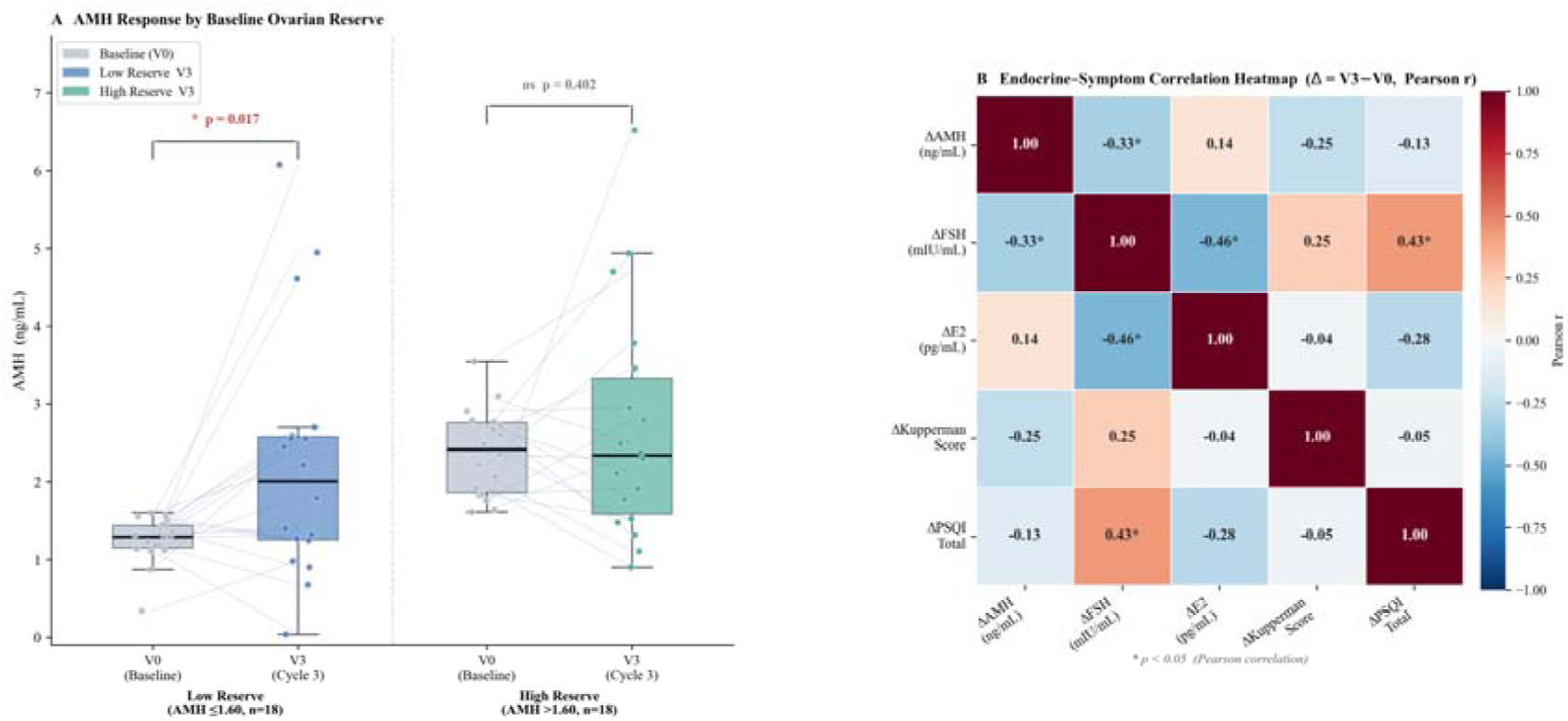
Subgroup and correlation analyses. (A) AMH at V0 and V3 stratified by baseline ovarian reserve (median AMH split), shown as box-and-whisker plots with jittered points. (B) Correlation heatmap of changes (Δ = V3 − V0) in hormonal (ΔAMH, ΔFSH, ΔE2) and clinical (ΔKupperman Index, ΔPSQI) measures; red = positive, blue = negative correlation, intensity proportional to Pearson r. *p < 0.05, **p < 0.01.

Correlation Matrix: Linking Hormones to Clinical Experience: To interrogate whether the improvements in sleep and menopausal symptoms were physiologically driven by EGT-induced hormonal shifts rather than placebo effects, a correlation heatmap was constructed (Figure 3B). We observed that the reduction in basal FSH (ΔFSH) was significantly correlated with the restoration of serum E2 (ΔE2, r = −0.46, p < 0.05) and AMH (ΔAMH, r = −0.33, p < 0.05). Furthermore, the improvement in sleep quality (reduced ΔPSQI) was significantly positively correlated with the reduction in FSH (r = 0.43, p < 0.05), mirroring the stabilization of the endocrine profile. While KI scores improved profoundly overall, their correlation with individual hormone shifts did not reach statistical significance. Nevertheless, the significant correlations surrounding FSH and sleep provide crucial mechanistic rationale, indicating that EGT-mediated microenvironmental stabilization in the ovary translates directly into systemic symptomatic relief.

### 3.4 Progressive Amelioration of Subjective Symptoms and Quality of Life

Continuous tracking of validated questionnaires across the four visits (V0 to V3) demonstrated progressive, time-dependent improvements across all subjective clinical domains (Figure 4):

**Figure 4.**
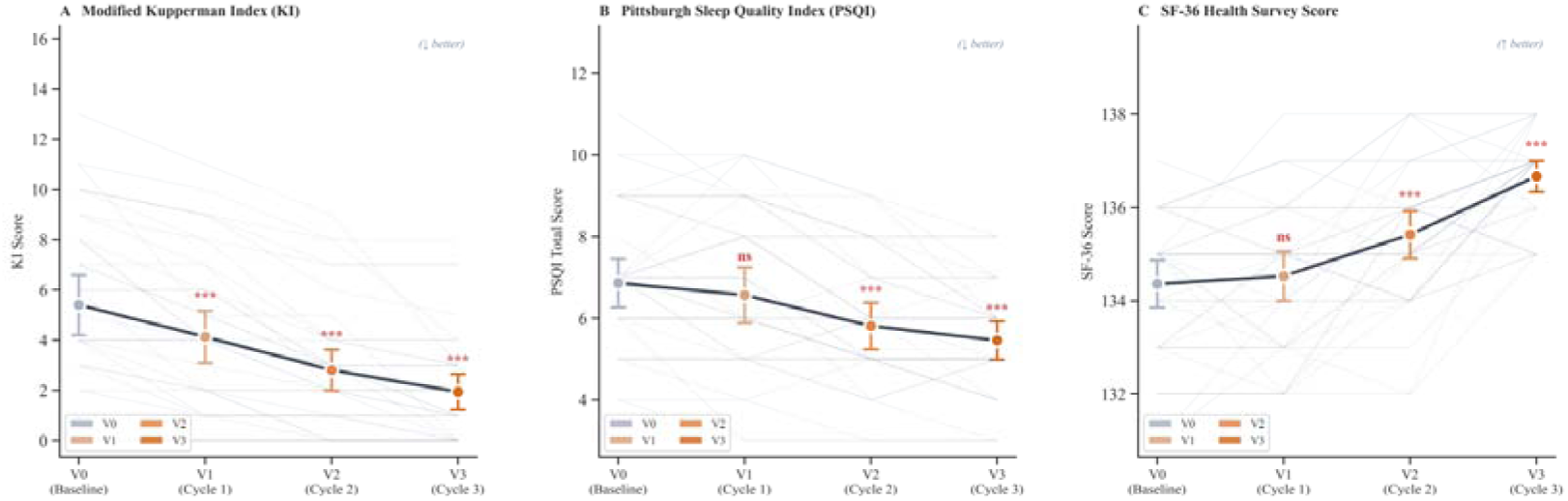
Time-course of subjective symptom and quality-of-life scores. Point plots show (A) modified Kupperman Index, (B) PSQI, and (C) SF-36 scores across four visits (V0–V3). Data are mean ± 95% CI; faint background lines trace individual trajectories (N = 36). ***p < 0.001 vs. baseline.

Reproductive Aging-Related Symptoms (KI): Modified KI scores decreased rapidly from 5.42 ± 3.66 at baseline to 3.50 ± 2.74 at the first cycle (V1) and stabilized at a low of 1.90 ± 2.16 by the third cycle (V3) (p < 0.0001) (Figure 4A).

Sleep Quality (PSQI): Sleep disturbances were progressively relieved, with PSQI scores dropping from 6.89 ± 1.82 to 5.50 ± 1.40 over the 3-month period (p < 0.0001). Individual spaghetti plots show that the vast majority of ‘poor sleepers’ (baseline PSQI > 5) responded positively to the intervention (Figure 4B).

Quality of Life (SF-36): Significant, sustained increases in overall quality of life scores were observed throughout the intervention, with mean SF-36 scores rising from 134.39 ± 1.50 at baseline to 136.70 ± 1.00 at V3 (p < 0.001) (Figure 4C).

#### Menstrual Characteristics

Detailed tracking of menstrual status revealed a distinct biphasic response across the intervention period (Figure 5). During the initial cycles (V1 and V2), menstrual scores exhibited a transient fluctuation. However, by the third cycle (V3), a robust physiological recovery was evident; the proportion of participants reporting optimal scores (score = 5) for cycle regularity, duration, volume, and pain progressively expanded, while the incidence of severe menstrual irregularities was substantially reduced.

**Figure 5.**
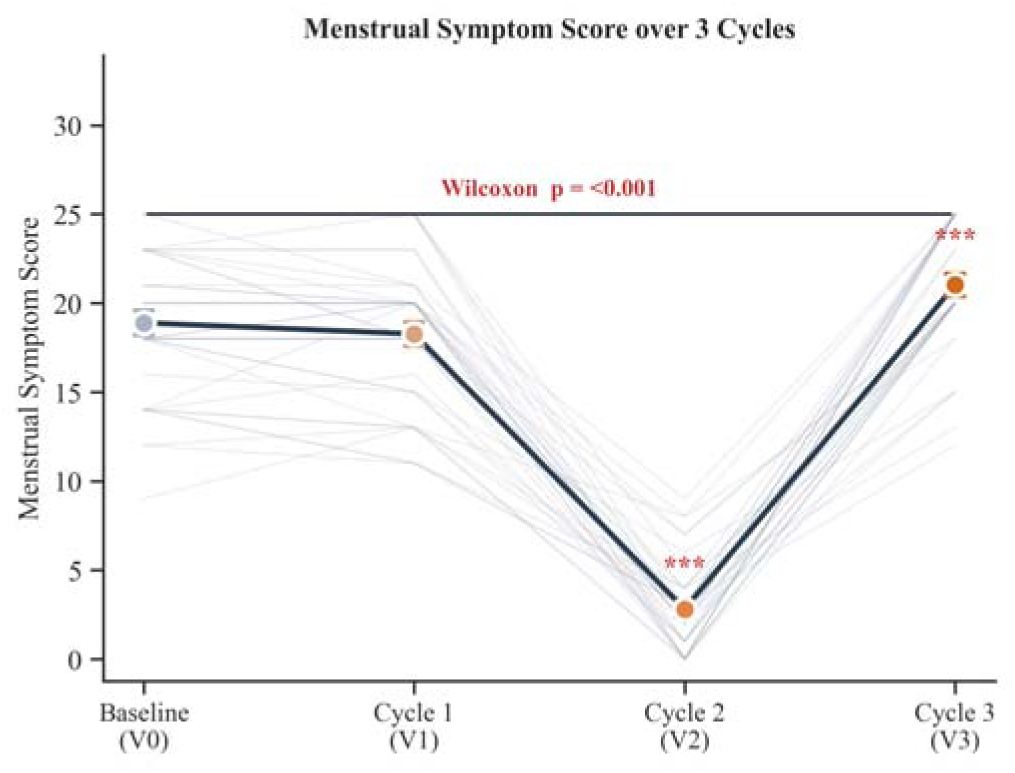
Menstrual characteristics over the 3-month intervention. Stacked bar charts show the proportional distribution of participant scores across five menstrual domains (cycle regularity, duration, volume, dysmenorrhea, predictability) at baseline (V0) and cycles 1–3 (V1–V3). Higher scores indicate better menstrual health.

### 3.5 Safety and Tolerability

The oral EGT intervention (120 mg/day) was exceptionally well-tolerated. No treatment-related adverse events (AEs) or dropouts due to side effects were reported. Comprehensive routine laboratory evaluations confirmed systemic safety. Core hematological indices, hepatic biomarkers, and renal function indicators largely remained highly stable without significant deviations from baseline (most p > 0.05). While blood urea nitrogen (BUN) exhibited a statistically significant decrease (p = 0.014), the values remained well within the healthy physiological range (2.6–7.5 mmol/L), indicating no clinical safety concerns (Supplementary Table S2).

## 4. Discussion

The decline of ovarian reserve is a central biological event that dictates the female reproductive lifespan and drives subsequent multi-systemic aging. This pilot study provides preliminary clinical observations that, in women with suboptimal ovarian reserve, oral EGT supplementation was associated with increases in ovarian reserve markers and improvements in associated symptoms over three menstrual cycles. As an open-label, single-arm study, it cannot establish causality, and the observed changes may partly reflect natural fluctuation, regression to the mean, or placebo effects. Following three menstrual cycles of continuous EGT supplementation, participants demonstrated a marked increase in serum AMH alongside a restorative shift in the basal FSH/E2 balance. Notably, the stratification by baseline ovarian reserve revealed a more pronounced AMH response in women with low baseline reserve (AMH ≤1.60 ng/mL; +0.99 ng/mL, p=0.017), while those with higher baseline reserve showed a smaller, non-significant change (+0.31 ng/mL, p=0.402). This suggests that EGT may offer greater benefit to those with a more depleted follicular pool, a finding that warrants prospective validation in larger cohorts.

A plausible mechanism is EGT’s capacity to scavenge intra-ovarian oxidative stress. Supporting this, a preclinical study demonstrated that L-ergothioneine supplementation reduces oxidative stress and apoptosis while improving the developmental competence and quality of in vitro–produced embryos [18]. However, as no oxidative-stress biomarkers were measured in the present study, this mechanism remains hypothetical and requires direct confirmation in our population. The cyclical maturation of ovarian follicles is an energy-intensive process highly reliant on mitochondrial oxidative phosphorylation, which inevitably leaks reactive oxygen species (ROS). In the aging ovary, ROS accumulation outpaces endogenous scavenging mechanisms, initiating lipid peroxidation that directly impairs granulosa cell function and AMH secretion [3, 4]. EGT, facilitated by its specific OCTN1 transporter, bypasses standard diffusion limitations to accumulate directly within the mitochondria of highly active or stressed cells [10]. By extinguishing ROS at their origin, EGT preserves the structural integrity of granulosa cells and interrupts the apoptotic signaling cascades that drive follicular atresia. This microenvironmental stabilization allows the surviving cohort of small antral follicles to function more efficiently, reflecting clinically as an elevation in AMH and an optimized FSH/E2 feedback loop.

Specifically, the significant reduction in elevated basal FSH levels serves as a direct consequence of restored ovarian function; as EGT improves the viable follicle pool, the concurrent rise in estradiol (E2) exerts a normalized negative feedback on the hypothalamic-pituitary axis, thereby suppressing FSH hypersecretion back to optimal ranges.

Beyond its peripheral action on the gonads, EGT exhibited profound effects on autonomic regulation and quality of life, as evidenced by the progressive decline in modified Kupperman and PSQI scores over the intervention period. Crucially, the statistical correlations between objective endocrine shifts (e.g., ΔE2, ΔAMH) and the amelioration of vasomotor symptoms (ΔKI) strongly support a physiological rationale for symptom relief rather than a mere placebo response. EGT may contribute to estradiol stabilization, which could in turn modulate hypothalamic thermoregulation, potentially alleviating vasomotor symptoms; however, this mechanistic pathway requires direct confirmation in future studies. Furthermore, EGT is a known neuroprotectant capable of crossing the blood-brain barrier. Its direct antioxidant action within the central nervous system may independently reduce neuroinflammation, lower sympathetic overdrive, and promote restorative sleep, thereby driving the comprehensive improvements observed in the SF-36 health survey.

Interestingly, the transient V-shaped fluctuation observed in menstrual scores during the early intervention phase (V1-V2) strongly aligns with the physiological timeline of folliculogenesis. Classic follicular dynamics models establish that the maturation from pre-antral to pre-ovulatory follicles requires approximately 85 days [19]. Consequently, follicles recruited and ovulated during the first two cycles had not yet fully benefited from continuous EGT tissue saturation and microenvironmental support restoration. Furthermore, the EGT-induced increase of circulating estradiol initiates a dynamic recalibration of the hypothalamic-pituitary-ovarian (HPO) axis [20]. This temporary endocrine readjustment may clinically manifest as transient menstrual irregularity before a new, optimized hormonal homeostasis is firmly established by Cycle 3 (Figure 5).

The safety profile of EGT is well established. High-purity EGT has received GRAS status from the U.S. FDA (GRN No. 1270) and a favorable novel-food safety assessment from EFSA, with toxicological studies indicating a wide safety margin (NOAEL well above the 120 mg/day dose used here). Prior human supplementation trials have reported excellent tolerability with no significant adverse effects, consistent with the absence of product-related adverse events in our cohort. [10–12].

This study has several strengths and limitations. Strengths include the use of objective biochemical and imaging markers and longitudinal follow-up across three cycles. Limitations include the open-label, single-arm design without a placebo control (precluding causal inference), the small sample size (N=36 completers) and exploratory nature, the increased variability of AMH at V3, the use of a non-validated menstrual questionnaire, and the single-center setting. These findings should therefore be regarded as hypothesis-generating, and a randomized, placebo-controlled trial is warranted.

## 5. Conclusion

In this open-label, single-arm pilot study, oral L-ergothioneine (120 mg/day) over three menstrual cycles was well tolerated and was associated with increases in serum AMH, favorable shifts in the FSH/E2 profile, and improvements in reproductive aging-related and sleep symptoms in women with suboptimal ovarian reserve. Owing to the absence of a control group, causality cannot be inferred. These preliminary, hypothesis-generating findings provide effect-size estimates to inform an adequately powered, randomized, placebo-controlled trial.

## Clinical Trial Registration

This trial was registered with the Chinese Clinical Trial Registry (Registration Number: ChiCTR2500104484) on June 18, 2025 (Prospectively registered).

## Ethics approval and consent to participate

This study was conducted in accordance with the Declaration of Helsinki and Good Clinical Practice (GCP) guidelines. The study protocol was formally approved by the Institutional Ethics Committee of Qingdao Central Hospital (Approval No.: SY202502202). Written informed consent to participate was obtained from all participants prior to screening.

## Data Availability Statement

The original data presented in this study are included in the article and its supplementary materials. De-identified individual participant data underlying the results reported in this article will be made available by the corresponding author upon reasonable request, subject to ethical and privacy considerations.

## Authors’ Contributions

G.X., W.D., and F.L. conceptualized and designed the study. C.G., and H.J. conducted the clinical trial and collected the data. J.C., W.L., and H.J. analyzed the data and performed the statistical analysis. W.L. wrote the first draft of the manuscript. All authors reviewed, edited, and approved the final manuscript.

## References

1. Broekmans FJ, Kwee J, Hendriks DJ, Mol BW, Lambalk CB. A systematic review of tests predicting ovarian reserve and IVF outcome. Hum Reprod Update. 2006;12(6):685–718. doi:10.1093/humupd/dml034

2. Harlow SD, Gass M, Hall JE, Lobo R, Maki P, Rebar RW, Sherman S, Sluss PM, de Villiers TJ; STRAW 10 Collaborative Group. Executive summary of the Stages of Reproductive Aging Workshop + 10: addressing the unfinished agenda of staging reproductive aging. Menopause. 2012 Apr;19(4):387–95. doi: 10.1097/gme.0b013e31824d8f40

3. Agarwal A, Aponte-Mellado A, Premkumar BJ, Shaman A, Gupta S. The effects of oxidative stress on female reproduction: a review. Reprod Biol Endocrinol. 2012; 10: 49. 10.1186/1477-7827-10-49

4. Lim J, Luderer U. Oxidative damage increases and antioxidant gene expression decreases with aging in the mouse ovary. Biol Reprod. 2011;84(4):775–782. 10.1095/biolreprod.110.088583

5. Sasaki H, Hamatani T, Kamijo S, et al. Impact of Oxidative Stress on Age-Associated Decline in Oocyte Developmental Competence. Front Endocrinol (Lausanne). 2019; 10: 811. 10.3389/fendo.2019.00811

6. La Marca A, Broekmans FJ, Volpe A, Fauser BC, Macklon NS; ESHRE Special Interest Group for Reproductive Endocrinology--AMH Round Table. Anti-Mullerian hormone (AMH): what do we still need to know? Hum Reprod. 2009;24(9):2264–2275. 10.1093/humrep/dep210

7. Cheah IK, Halliwell B. Ergothioneine; antioxidant potential, physiological function and role in disease. Biochim Biophys Acta. 2012;1822(5):784–793. 10.1016/j.bbadis.2011.09.017

8. Gründemann D. The ergothioneine transporter controls and indicates ergothioneine activity—A review. Prev Med. 2012; 54: S71–S74. 10.1016/j.ypmed.2011.12.001

9. Paul BD, Snyder SH. The unusual amino acid L-ergothioneine is a physiologic cytoprotectant. Cell Death Differ. 2010;17(7):1134–1140. 10.1038/cdd.2009.163

10. Liu H, et al. Ergothioneine as a Natural Antioxidant Against Oxidative Stress-Related Diseases. Front Pharmacol. 2022; 13: 850813. 10.3389/fphar.2022.850813

11. Marone PA, Trampota J, Weisman S. A Safety Evaluation of a Nature-Identical l-Ergothioneine in Sprague Dawley Rats. Int J Toxicol. 2016;35(5):568–583. 10.1177/1091581816653375

12. Turck D, Bresson JL, Burlingame B, et al. Statement on the safety of synthetic l-ergothioneine as a novel food - supplementary dietary exposure and safety assessment for infants and young children, pregnant and breastfeeding women. EFSA J. 2017;15(11): e05060.

13. GRN No. 1270 L-ergothioneine produced by Escherichia coli K-12 MG1655 expressing enzymes from Neurospora crassa and Mycolicibacterium smegmatis MC2 155 [EB/OL]. (2026-01-22) [2026-03-16]. https://hfpappexternal.fda.gov/scripts/fdcc/index.cfm?set=GRASNotices&id=1270

14. Tang RMY, Cheah IK, Yew TSK, Halliwell B. Distribution and accumulation of dietary ergothioneine and its metabolites in mouse tissues. Sci Rep. 2018;8(1):1601. 10.1038/s41598-018-20021

15. Kupperman HS, Blatt MH, Wiesbader H, Filler W. Comparative clinical evaluation of estrogenic preparations by the menopausal and amenorrheal indices. J Clin Endocrinol Metab. 1953;13(6):688–703. 10.1210/jcem-13-6-688

16. Buysse DJ, Reynolds CF 3rd, Monk TH, Berman SR, Kupfer DJ. The Pittsburgh Sleep Quality Index: a new instrument for psychiatric practice and research. Psychiatry Res. 1989;28(2):193–213. 10.1016/0165-1781(89)90047-4

17. Ware JE Jr, Sherbourne CD. The MOS 36-item short-form health survey (SF-36). I. Conceptual framework and item selection. Med Care. 1992;30(6):473–483.

18. Zullo G, Albero G, Neglia G, De Canditiis C, Bifulco G, Campanile G, Gasparrini B. L-Ergothioneine supplementation during culture improves quality of bovine in vitro-produced embryos. Theriogenology. 2016;85(4):688–697. 10.1016/j.theriogenology.2015.10.006

19. Gougeon, A. “Dynamics of follicular growth in the human: a model from preliminary results.” Human reproduction (Oxford, England) vol. 1,2 (1986): 81–7. 10.1093/oxfordjournals.humrep.a136365

20. Richards, Joanne S, and Stephanie A Pangas. “The ovary: basic biology and clinical implications.” The Journal of clinical investigation vol. 120,4 (2010): 963–72. 10.1172/JCI41350

