## Supplementary_Materials for "Oral Gene III^®^ L-Ergothioneine Capsules in Women with Suboptimal Ovarian Reserve: A Single-Center, Open-Label, Self-Controlled Pilot Study of Ovarian Reserve Markers, Endocrine Profiles, and Reproductive Aging-Related Symptoms" (from another version of this preprint: version 1, 2026-04-04)

**Figure S1. Individual trajectories and distribution of Antral Follicle Count (AFC).**

Violin plots overlaid with semi-transparent spaghetti lines illustrate the distribution and individual trajectories of AFC from baseline (V0) to Cycle 3 (V3). While strictly not statistically significant (p = 0.4315), a visible upward trend in the growing follicle pool was observed.

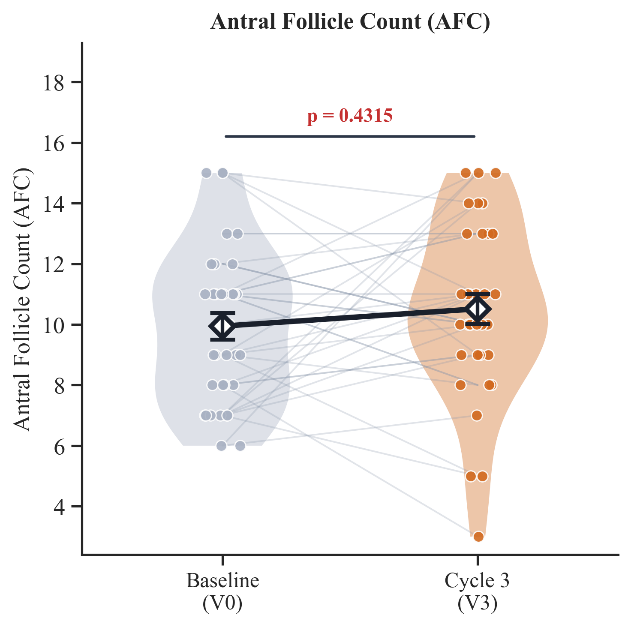

**Table S1. Exploratory Secondary Endpoints (V0 vs. V3)**

Data are presented as Mean ± SD. p-values were calculated using paired t-tests or Wilcoxon signed-rank tests depending on data normality.

| **Variable** | **Baseline (V0)**  **Mean ± SD** | **Cycle 3 (V3)**  **Mean ± SD** | **p value** |
| --- | --- | --- | --- |
| Antral Follicle Count (AFC)  (count)(n=35) | 10.1 ± 2.6 | 10.8 ± 3.20 | 0.432 |
| Luteinising Hormone (LH)  (mIU/mL) (n=36) | 5.87 ± 2.12 | 6.24 ± 2.87 | 0.963 |
| Prolactin (PRL)  (mIU/L) (n=36) | 341.90 ± 183.82 | 394.91 ± 298.23 | 0.487 |
| Testosterone (T)  (ng/mL) (n=36) | 0.24 ± 0.12 | 0.22 ± 0.11 | 0.144 |
| Progesterone (P)  (ng/mL) (n=36) | 0.66 ± 2.5 | 1.63 ± 5.14 | 0.220 |

**Table S2. Safety and Tolerability Profile**

Data are presented as Mean ± SD. Routine hematological and biochemical markers showed no significant clinical alterations.

| **Variable** | **Baseline (V0)**  **Mean ± SD** | **Cycle 3 (V3)**  **Mean ± SD** | **p value** |
| --- | --- | --- | --- |
| **Liver Function** | | | |
| ALT (U/L) (n=36) | 16.83 ± 7.56 | 16.83 ± 6.54 | 0.886 |
| AST (U/L) (n=36) | 19.67 ± 4.83 | 20.17 ± 4.56 | 0.635 |
| Total Bilirubin  (μmol/L) (n=36) | 9.92 ± 3.87 | 9.99 ± 3.94 | 0.377 |
| ALP (U/L) (n=36) | 64.19 ± 12.42 | 65.31 ± 13.28 | 0.760 |
| GGT (U/L) (n=36) | 13.42 ± 8.21 | 13.83 ± 12.10 | 0.590 |
| **Renal Function** | | | |
| BUN (mmol/L) (n=36) | 4.57 ± 0.77 | 4.07 ± 0.89 | 0.014 * |
| Creatinine (μmol/L) (n=36) | 51.25 ± 6.23 | 49.06 ± 6.38 | 0.103 |
| eGFR  (mL/min/1.73m²) (n=36) | 118.33 ± 3.70 | 119.44 ± 4.37 | 0.121 |
| **Haematology** | | | |
| WBC (×10⁹/L) (n=36) | 5.89 ± 1.37 | 5.95 ± 1.57 | 0.780 |
| RBC (×10¹²/L) (n=36) | 4.30 ± 0.35 | 4.26 ± 0.30 | 0.337 |
| Haemoglobin (g/L) (n=36) | 125.25 ± 11.64 | 125.28 ± 11.27 | 0.938 |
| Platelets (×10⁹/L) (n=36) | 255.07 ± 58.80 | 249.17 ± 55.80 | 0.440 |
